# Comparison of EMG, Video, and Actigraphy Signals for Detecting Motor Activity in REM Sleep Behavior Disorder

**DOI:** 10.64898/2026.02.18.26346544

**Authors:** Kang Hyun Ryu, Giorgio Ricciardiello Mejia, Salonee Marwaha, Andreas Brink-Kjaer, Emmanuel H. During

## Abstract

**Background/Objectives:** Electromyography (EMG), video-polysomnography (vPSG), and wrist actigraphy are each used to develop diagnostic algorithms for Rapid eye movement sleep behavior disorder (RBD). However, the extent to which they capture overlapping versus distinct motor phenomena remains unknown. We evaluated the respective contributions of actigraphy, EMG and vPSG to the measurement of REM-sleep motor activity.

**Methods:** Seventeen adults with RBD (Mount Sinai n = 9; Stanford n = 8) and eight control participants from an open Newcastle dataset underwent vPSG and concomitant wrist actigraphy. Flexor digitorum superficialis EMG activity and video-detected movements were manually scored in 3-second mini epochs. Actigraphy was quantified using an acceleration-magnitude–based activity count model. Statistical and agreement analyses were performed to assess the motor events captured by all three, any two, or by each modality independently during REM sleep.

**Results:** In participants with RBD, actigraphy-derived movement load was significantly higher during REM sleep than during non-REM stages, a pattern not observed in control participants. Across 12,941 3-second mini epochs, EMG, actigraphy, and video detected 1,703, 1,613, and 811 motor events, of which 413 were detected concurrently by all three modalities. Pairwise agreement was moderate and increased from EMG–actigraphy (κ = 0.27 ± 0.10) to actigraphy–video (κ = 0.41 ± 0.12) and EMG–video (κ = 0.45 ± 0.15). Of EMG-detected events, 49.0% were also detected by actigraphy; of actigraphy-detected events, 37.2% were detected by EMG and 34.9% by video. Actigraphy activity counts were highest for events detected by all three modalities and lowest for actigraphy-only events.

**Conclusion:** Actigraphy-measured REM-related motor activity was elevated in RBD but not in controls. EMG, actigraphy, and video captured partially overlapping motor events in RBD patient, with actigraphy showing the highest sensitivity and manually scored video the lowest.

## Introduction

Rapid-eye-movement sleep behavior disorder (RBD) is a parasomnia characterized by the loss of normal muscle atonia during REM sleep, resulting in motor disinhibition^1^. Motor manifestations in RBD vary widely in intensity both across individuals and from night to night, ranging from subtle twitches to complex and occasionally violent behaviors. Prompt and accurate diagnosis of RBD is critical, as it is most often a prodromal manifestation of synucleinopathies, including Parkinson’s disease, dementia with Lewy bodies, and multiple system atrophy.

The diagnosis of RBD relies on overnight video-polysomnography (vPSG). This assessment includes electromyography (EMG) recordings from the submentalis (chin), flexor digitorum superficialis (upper extremities), and anterior tibialis muscles (lower extremities) to quantify muscle activity during REM sleep, along with synchronized infrared video recording, as outlined by International RBD Study Group guidelines^2^. EMG provides a direct window into the pathophysiologic hallmark of RBD— excessive muscle activity during REM sleep, whereas video data are particularly valuable for evaluating RBD symptom severity^3,4^.

Importantly, EMG activity does not always correspond to overt movements, as some electrical activity may not reach a movement threshold. Conversely, not all movements detected on video are captured by EMG, as they may involve muscle groups that are not monitored. Yet, movements in video may be obscured by bed coverings, and subtle twitches may fall below the detection threshold of infrared cameras. Recent studies have explored the potential of video analysis for differentiating RBD from other sleep disorders^5,6.^

A third modality, wrist actigraphy, has been shown to capture excessive movements in RBD compared to other sleep disorders and healthy sleep^7,8,9^. Despite a growing body of evidence supporting the development of actigraphy-based classifiers—and the potential scalability of this approach as a screening tool—two critical aspects of wrist accelerometry in RBD remain insufficiently studied: (1) the extent to which movements detected in RBD are specific to REM sleep, as opposed to non-REM sleep or wakefulness; and (2) the degree to which wrist actigraphy captures overlapping versus distinct motor phenomena of REM sleep when compared with EMG–particularly of the upper extremities, and with movements observed on video.

## Methods

### 2.1 Participants

Three research cohorts were used in this study:

a. RBD cases. 17 adult participants with RBD (n = 9) at Mount Sinai Sleep Research Center, (n = 8) at the Stanford Sleep Research Center; (n = 16) with idiopathic RBD, (n = 1) with antidepressant-induced RBD underwent vPSG with concomitant wrist actigraphy between June 2021 and November 2025. All RBD diagnoses were confirmed by overnight vPSG based on ICSD-3 criteria^10^. Research participants signed an informed consent form, and the study was approved by the Institutional Review Boards of the Icahn School of Medicine at Mount Sinai. Participant demographics are summarized in **Table 1** below.
b. Controls. The open research Newcastle dataset^11^ (n = 8), in which participants also wore wrist actigraphy during concomitant vPSG, was used for this study.

**Table 1.** Participant demographics and clinical characteristics. Values are reported as mean ± standard deviation or number of participants (n), as indicated. iRBD = idiopathic REM sleep behavior disorder; OSA = obstructive sleep apnea, as defined by ≥ 5 apneas and/or hypopneas resulting in ≥ 4% oxygen desaturations per hour of sleep; RLS = restless legs syndrome; PLM = periodic limb movements, as defined by a PLM index ≥ 15 per hour of sleep.

|  | Mount Sinai RBD<br>Participants (n = 9) | Stanford RBD<br>participants (n = 8) | Control participants<br>(n = 9) |
| --- | --- | --- | --- |
| Age (years) | 57.5 ± 14.3 | 65.6 ± 8.23 | 49.4 ± 12.8 |
| Sex (male, n) | 9 | 7 | 7 |
| BMI (kg/m<sup>2</sup>) | 26.3 ± 2.86 | 28.5 ± 5.95 | N/A |
| iRBD (n) | 8 | 8 | 0 |
| antidepressant-induced<br>RBD (n) | 1 | 0 | 0 |
| Years from symptom onset | 5 ± 6.09* | N/A | N/A |
| Melatonin (n) | 2 | 4 | N/A |
| Clonazepam (n) | 0 | 3 | N/A |
| OSA (n) | 3 | 0 | 1 |
| RLS (n) | 2 | 0 | 0 |
| PLM (n) | 1 | 2 | N/A |

\*n=8

The Mount Sinai RBD cohort is the primary dataset in this study and the only one with complete vPSG and bilateral actigraphy recordings available for the comparison of each modality. The Stanford RBD and Newcastle control cohorts were included in the initial analysis reporting motor activity across sleep stages in order to increase the sample size in RBD and provide data from healthy controls.

### 2.2 Video PSG

All RBD participants were connected to PSG equipment 2 hours prior to their habitual bedtime, which was determined by participant interview. Data (diagnostic or on a therapeutic Continuous Positive Airway Pressure (CPAP) machine) was acquired following standard American Academy of Sleep Medicine (AASM) protocols using Compumedics E-series and Grael 2 systems (Melbourne, Australia). Signal acquisition included 19 channel electroencephalography placed according to the International 10– 20 system or a subset of 6 channels (F3, F4, C3, C4, O1, and O2), left and right electro-oculography (EOG), all sampled at 256 or 512 Hz and referenced to a common patient and ground electrode, submental bipolar, bilateral flexor digitorum superficialis, and anterior tibialis bipolar EMG, respiration by a pressure transducer and PAP device interface, effort by rib/abdomen impedance plethysmography, single-channel ECG, and oxygen saturation by pulse oximetry (SpO_2_). Synchronized infrared video was recorded continuously throughout the night.

Sleep stages were scored by a sleep technician in 30-s epochs according to standard criteria for sleep and EEG arousals and confirmed by a board-certified sleep specialist. Total sleep time (TST) and percentage time spent in wake, REM sleep, non-REM stage 1 (N1), non-REM stage 2 (N2), and Slow-Wave Sleep (SWS/N3) were determined.

A summary of the Mount Sinai, Stanford, and Newcastle vPSG data analyzed in this study is presented in **Tables 2, S1, and S2**. REM sleep periods shorter than 5 minutes were excluded from analysis, in line with International RBD Study Group guidelines^2^. Consequently, one REM period was excluded for Mount Sinai participant 6 (leaving three REM periods) and for control participants 1 and 2 (leaving two REM periods each).

**Table 2.** Sleep architecture characteristics for each RBD participant in the Mount Sinai cohort. Total sleep time, total REM sleep duration (with percentage of total sleep time), and number of REM sleep periods during the recorded overnight video-polysomnography (vPSG) are shown for each participant.

| Mount Sinai RBD Participant | Total sleep time (minutes) | Total REM duration (minutes) | REM sleep periods (counts) |
| --- | --- | --- | --- |
| 1 | 273 | 51.55 (18.9%) | 3 |
| 2 | 326 | 93.5 (28.7%) | 2 |
| 3 | 312 | 42.55 (13.6%) | 3 |
| 4 | 487.5 | 83.55 (17.1%) | 3 |
| 5 | 413 | 97.85 (23.7%) | 2 |
| 6 | 410.5 | 45 (11.0%) | 3 |
| 7 | 404.5 | 83.5 (20.6%) | 4 |
| 8 | 521.5 | 111.55 (21.4%) | 4 |
| 9 | 392 | 38 (9.69%) | 1 |

### 2.3 Actigraphy

All RBD participants wore an AX6 triaxial accelerometer (Axivity Ltd., Newcastle, UK) on both wrists during their overnight vPSG, recorded at 50 Hz with a ± 8 g dynamic range. Timestamps across EMG, infrared video, and actigraphy were synchronized by aligning the prominent movement artifacts generated by a standardized brisk arm maneuver performed at the start of the vPSG recording. An illustrative example of the study design is shown in **Figure 1**.

**Figure 1.**
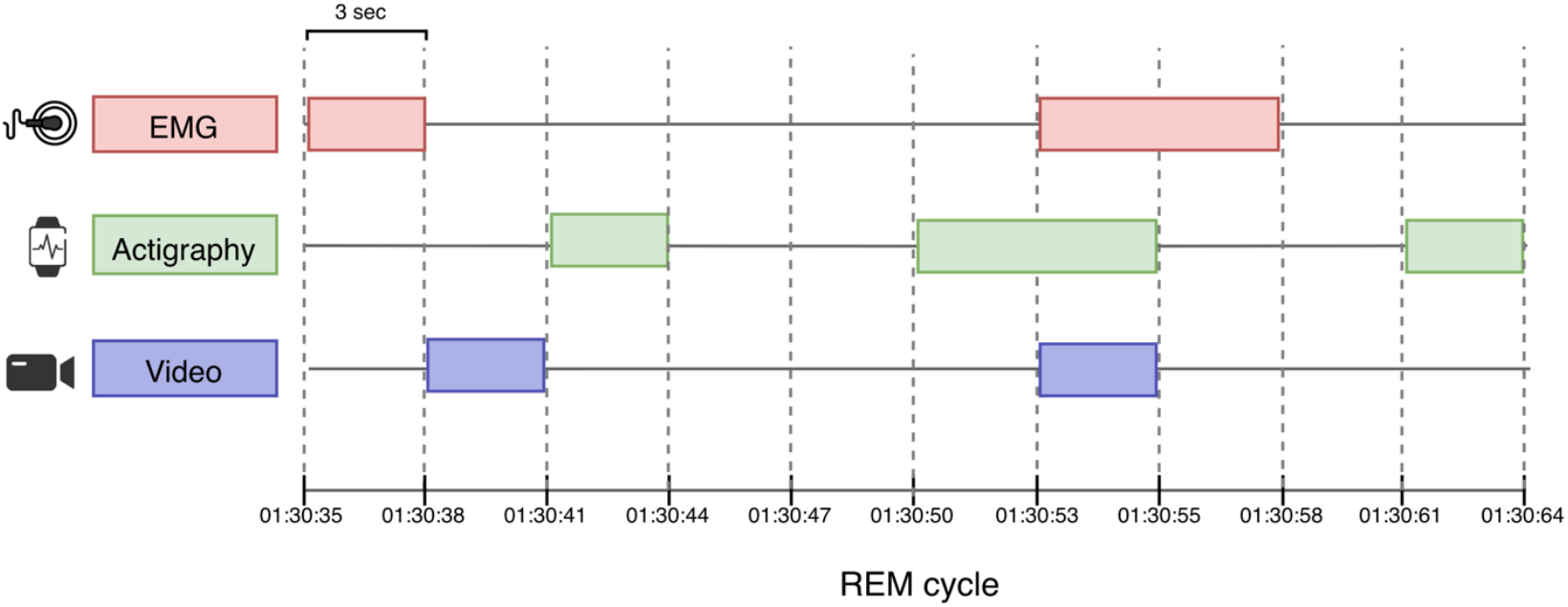
Study design and epoch-level synchronization. Synchronized bilateral flexor digitorum superficialis EMG, wrist actigraphy, and infrared video were recorded during overnight video polysomnography (vPSG) at the Mount Sinai Sleep Research Center. Motor activity detected by each modality was scored in contiguous 3-second mini-epochs during REM sleep. Colored blocks indicate sample active epochs for EMG (red), actigraphy (green), and video (blue).

All control participants in the Newcastle dataset wore a GENEActive actigraph^11^ (Activinsights Ltd., Kimbolton, UK).

### 2.4 Electromyographic scoring

Bilateral flexor digitorum superficialis EMG activity during REM sleep was scored according to the AASM Manual for the Scoring of Sleep and Associated Events, Version 3^12^. EMG signals were filtered using a 5 Hz high-pass filter and a 500 Hz low-pass filter. Within each REM sleep period, EMG activity was manually scored in 3-second mini-epochs using a binary system. Activity was defined as an amplitude at least twice the participant’s baseline level during that REM sleep cycle and lasting between 0.1 and 5.0 seconds.

### 2.5 Video scoring

Arm movements during each REM sleep cycle were manually evaluated from the video recordings and scored using a binary system. Video scoring was performed blinded to the EMG and actigraphy scores to minimize bias. Any observable arm movement within each 3-second mini-epoch was marked as activity. Movements were not differentiated by amplitude; small twitches and large, full-body movements were treated equivalently and coded as 1 (activity).

### 2.6 Actigraphy scoring: activity counts

Activity counts from the accelerometer data were calculated following the activity count model described in Brink-Kjaer et al. (2022)^7^. In brief, raw accelerometer data was calibrated such that the vector magnitude is equal to the gravitational acceleration (1 g) during periods of inactivity. Activity counts were calculated as the acceleration magnitude based on the Euclidean Norm Minus One formula:

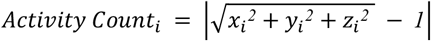

where *x*_*i*_, *y*_*i*_, and *z*_*i*_ represent acceleration along the three orthogonal axes. The activity counts were then averaged in 1-second bins. To mitigate baseline drift, the resulting time series were forward-backward filtered using a high-pass infinite impulse response (IIR) Butterworth filter with a stopband attenuation of 80 dB at 0.1 Hz and passband ripple of 1 dB starting at 0.5 Hz. Activity counts less than 0.1 were rounded down to zero. A 3-second mini epoch was classified as active if the maximum activity count within the epoch exceeded zero.

Accelerometer data from control participants in the Newcastle dataset were processed using a modified version of the open-dataset processing code^11,13^, and activity counts were calculated using the same algorithm described above.

### 2.7 Statistical and agreement analysis

Differences in actigraphy-derived activity counts across sleep stages were first examined to evaluate whether detected movements were specific to REM sleep rather than non-REM sleep and wakefulness, and whether REM motor activity was elevated in participants with RBD relative to controls. We quantified actigraphy-derived movement load, defined as the sum of left and right wrist activity counts per minute, for each sleep stage. Within-participant pairwise comparisons of movement load between sleep stages were conducted using Wilcoxon signed-rank tests, with Holm p value correction for multiple comparisons.

Because motor activity-event prevalence differed across participants and modalities, we prioritized agreement metrics that are more robust to marginal imbalance. Cross-modality agreement was quantified using Cohen’s κ (chance-corrected agreement)^14^. Using EMG as the reference standard, we additionally computed sensitivity, specificity, and positive predictive value (PPV) for actigraphy and video. We also calculated conditional probabilities (P(Actigraphy=1∣Video=1) and P(Video=1|Actigrpahy=1)) between actigraphy and video to characterize their co-detection patterns. In addition, modality-detected motor activity rates and ratios were compared to quantify the proportion of activity captured by each modality. All metrics were computed per participant and then summarized across participants.

Finally, to assess whether actigraphy movement magnitude varied with multimodal consensus, we compared accelerometer-derived activity counts across mutually exclusive multimodal agreement categories (actigraphy alone; actigraphy and video; EMG and actigraphy; and all three modalities). Specifically, REM mini epochs were stratified by overlapping detection condition, and for each category we computed the mean actigraphy activity count across epochs. These values were then compared across conditions to assess how actigraphy-detected movement intensity changed as a function of modality co-detection.

## Results

### 3.1 Motor activity during REM vs NREM

Bilateral wrist actigraphy activity counts demonstrated sleep–wake state–dependent variability in participants with RBD (**Figure 2; Figures S1-S2**). Activity was highest in mean amplitude [0.233 (0.113)] and frequency [14.0 (2.77)] during wake and was generally lower and sparser across NREM sleep (amplitude [0.027 (0.013)], frequency [2.88 (1.40)]). REM epochs exhibited intermittent clusters of elevated activity relative to NREM.

**Figure 2.**
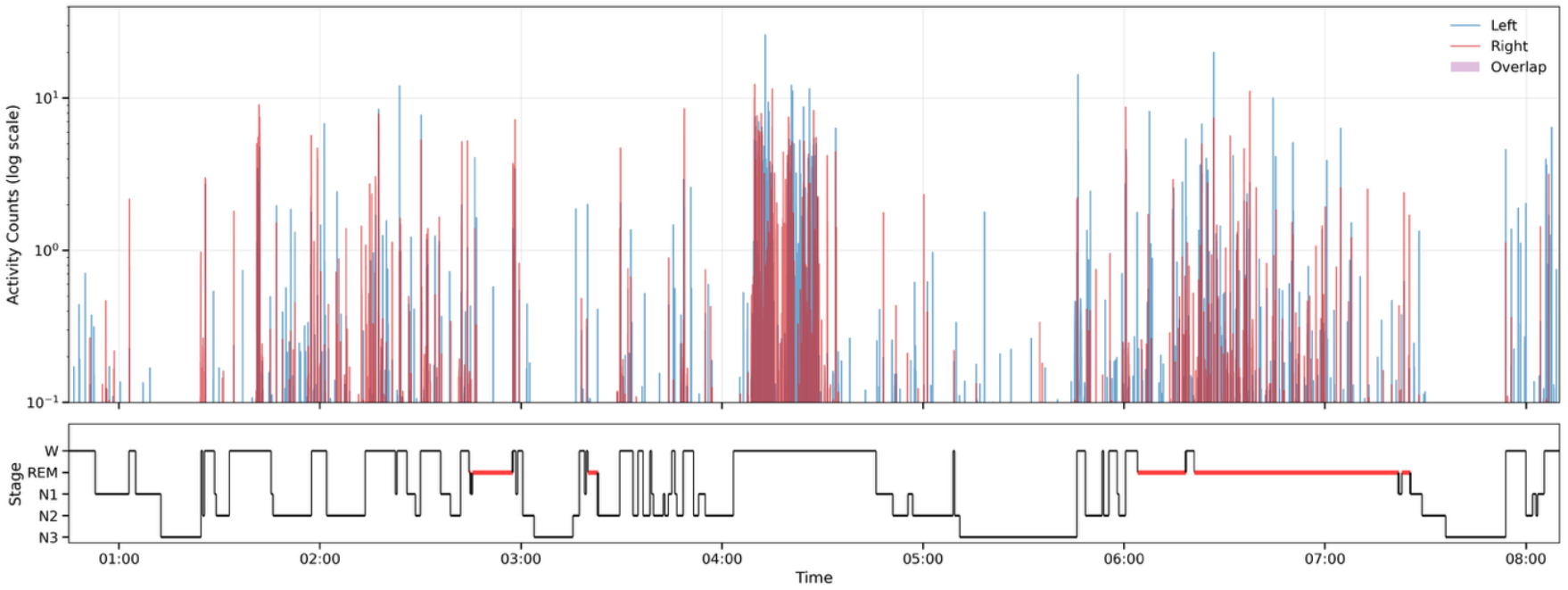
Sample bilateral wrist actigraphy aligned to sleep stages in an RBD participant (Mount Sinai participant 2). The upper panel shows left-(blue) and right-wrist (red) activity counts (log scale) across the overnight recording; purple shading marks epochs with concurrent activity in both wrists (overlap). The lower panel shows the corresponding hypnogram (W, N1, N2, N3, REM), with REM periods highlighted in red. Plots for the remaining Mount Sinai participants are provided in Supplementary Figures S2–S3. *Note*. The y-axis is truncated at 15 to improve readability.

In control participants, the highest actigraphy movement load was found in N1 (median 0.94 [IQR 1.97]), and lower across deeper stages of sleep, i.e., N2 (median 1.13 [IQR 0.56]), N3 (median 0.81 [IQR 1.05]), and REM sleep (median 0.90 [IQR 1.04]) (see **Figure 3**). No significant differences were observed between the NREM and REM stages. In contrast, in RBD participants, significant increase was observed in REM relative to NREM sleep. The movement load was highest during REM sleep (median 2.91 [IQR 2.20]), followed by N1 (median 1.72 [IQR 2.99]), N2 (median 0.80 [IQR 0.97]), and N3 sleep (median 0.42 [IQR 0.51]) with statistically significant differences in REM sleep compared to N2 (adjusted p = 0.003) and N3 sleep (adjusted p = 0.019). Movement load during REM sleep was higher in RBD participants than in controls (Mann–Whitney U test, uncorrected p = 0.046), with a large effect size (Cliff’s δ = 0.49). However, this effect did not remain significant after false discovery rate correction across sleep stages (adjusted p = 0.185).

**Figure 3.**
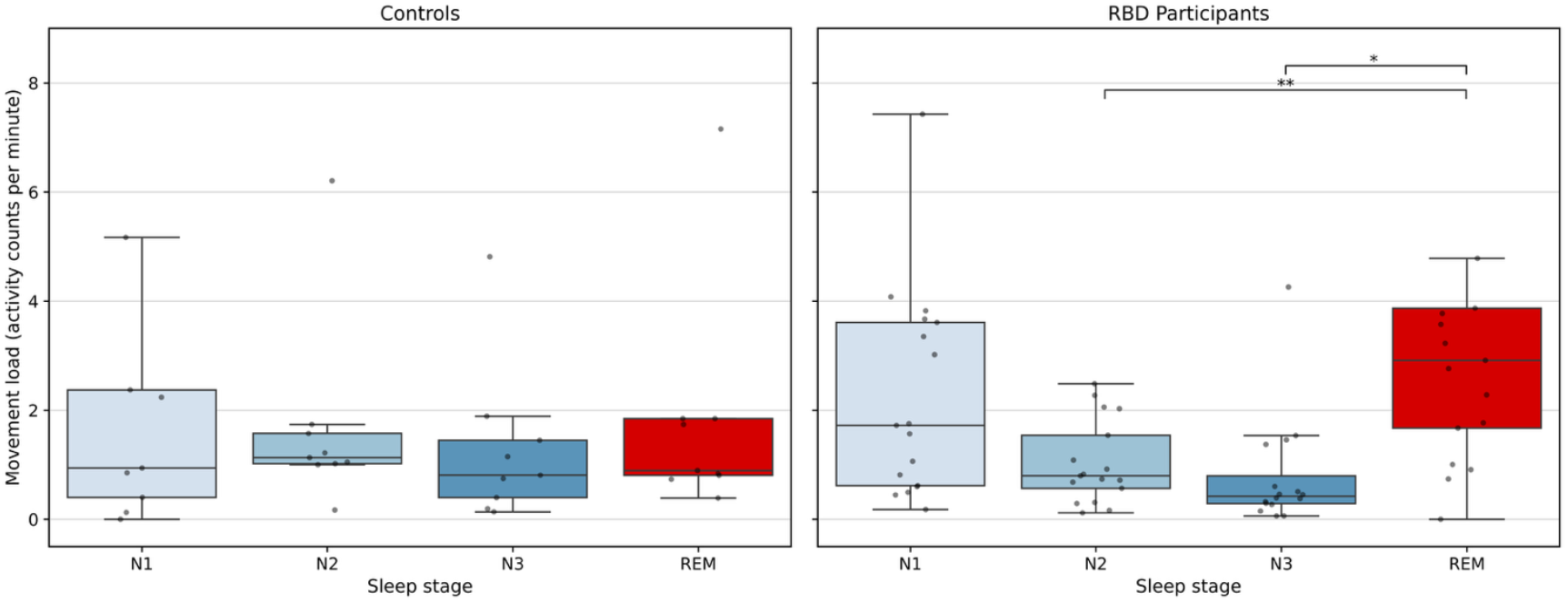
Movement load across sleep stages in RBD (top) and control participants (bottom). Movement load, defined as the sum of left and right wrist actigraphy activity counts per minute, is shown for N1, N2, N3, and REM (R) sleep. Each dot represents a single participant, and boxplots summarize the distribution across participants. Pairwise comparisons were conducted using Wilcoxon signed-rank tests with Holm correction for multiple comparisons. For RBD patients, movement load during REM sleep was significantly greater than during N2 (adjusted p = 0.006) and N3 sleep (adjusted p = 0.04). *Note*. Some higher values (9.69 for controls, N1; 9.21, 9.22, 10.2 for RBD participants, REM) are not shown to improve scale and readability.

### 3.2 Comparison of motor activity detected by EMG, actigraphy and video

Across 12941 3-second REM mini-epochs recorded across participants, 2797 contained motor events detected on the left, right, or both sides—captured by one, two, or all modalities. Of these, EMG, wrist actigraphy, and video detected 1703, 1613, and 811 motor events, respectively (**Figure 4**). Of these motor events, 413 events were detected simultaneously by all three modalities. 174 events were detected by both EMG and actigraphy, 187 events were detected by EMG and video, and 143 events were detected by actigraphy and video. Modality-specific detections were also observed, with 929 events detected by EMG alone, 883 by actigraphy alone, and 68 by video alone.

**Figure 4.**
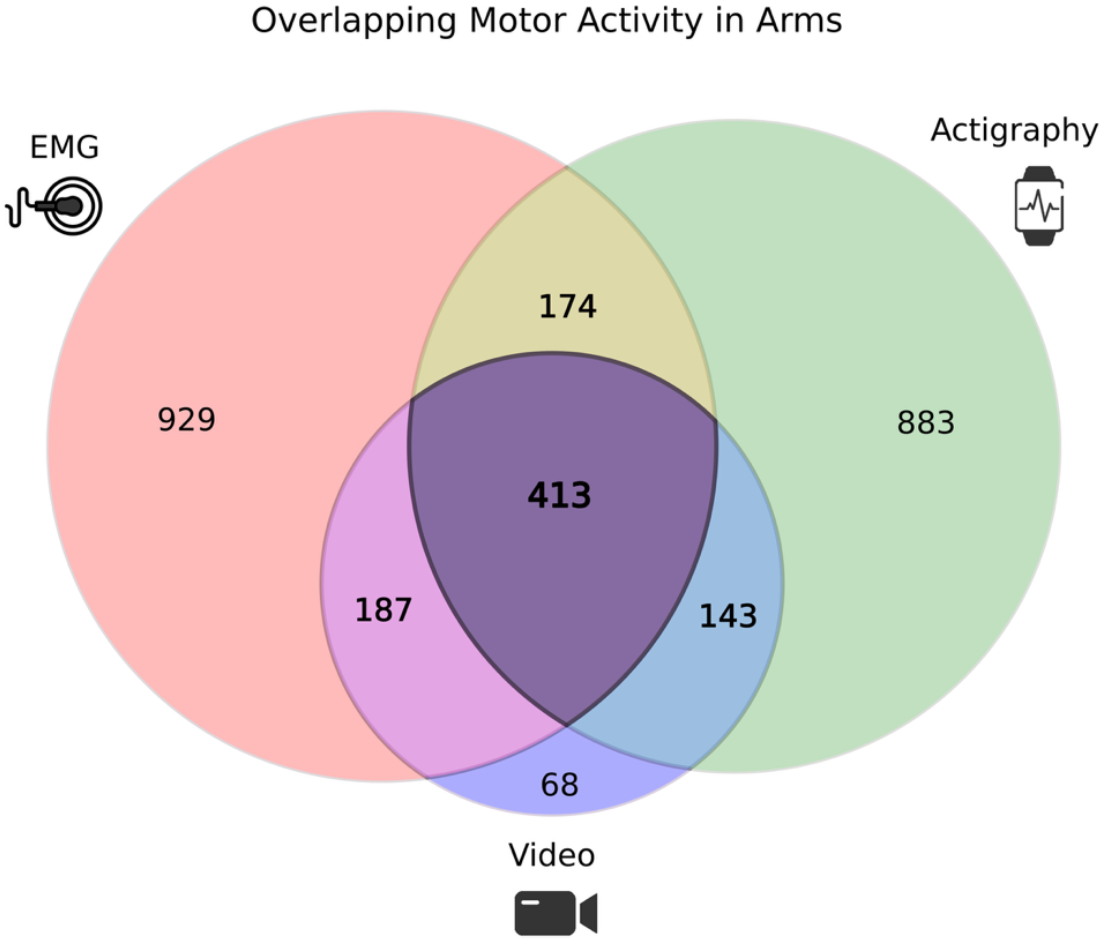
Overlap of REM-related motor activity detected by EMG, actigraphy, and video. This diagram illustrates, among the 12,941 three-second REM mini-epochs recorded across all participants, the number of motor events detected by EMG, wrist actigraphy, and infrared video on the left, right, or both sides of the arm. Values indicate the number of events detected uniquely by each modality or concurrently by two or all three modalities.

Agreement between modalities during REM sleep was assessed using Cohen’s κ (**Table 3; Figure S3**). Overall, pairwise agreement was moderate, with mean κ values increasing from EMG– actigraphy (κ = 0.27 ± 0.10) to actigraphy–video (κ = 0.41 ± 0.12) and EMG–video (κ = 0.45 ± 0.15). Consistent with these results, odds ratio analyses (log scale) showed the strongest association between EMG-video (pooled OR ≈ 49; 95% CI 29.8–80.3), followed by actigraphy-video (pooled OR ≈ 33; 95% CI 16.6–66.9) (**Figure S4**). EMG–actigraphy exhibited the weakest, but still robust and positive association (pooled OR ≈ 10.6; 95% CI 6.26–17.9).

**Table 3.** Agreement between EMG, actigraphy, and video detections during REM sleep. Cohen’s κ values are shown for pairwise comparisons between EMG, actigraphy, and video for each participant. Mean and standard deviation across participants are reported.

| Mount Sinai<br>RBD<br>Participant | Cohen's $\kappa$ | | |
| --- | --- | --- | --- |
|  | EMG vs. Acti | EMG vs. Video | Acti vs. Video |
| 1 | 0.25 | 0.58 | 0.34 |
| 2 | 0.08 | 0.28 | 0.21 |
| 3 | 0.40 | 0.41 | 0.34 |
| 4 | 0.34 | 0.27 | 0.60 |
| 5 | 0.27 | 0.32 | 0.48 |
| 6 | 0.39 | 0.59 | 0.54 |
| 7 | 0.24 | 0.58 | 0.39 |
| 8 | 0.19 | 0.39 | 0.43 |
| 9 | 0.27 | 0.63 | 0.36 |
| Mean | 0.27 | 0.45 | 0.41 |
| Std Dev | 0.10 | 0.14 | 0.12 |

Using EMG as the reference standard, we quantified the diagnostic performance of wrist actigraphy and infrared video for detecting REM-related motor activity (**Table 4**). Across participants, actigraphy demonstrated slightly higher sensitivity to EMG-defined motor events than video (mean ± SD: 0.49 ± 0.19 vs. 0.46 ± 0.19), indicating that actigraphy captured a greater proportion of motor events detected by EMG. Both modalities exhibited high specificity relative to EMG, although specificity was higher for video than for actigraphy (0.98 ± 0.01 vs. 0.90 ± 0.06). PPV differed markedly between modalities: video detections were more likely to correspond to concurrent EMG activity (PPV = 0.67 ± 0.22), whereas actigraphy detections showed lower PPV (0.37 ± 0.21), reflecting a higher proportion of actigraphy-only detections in the absence of EMG-defined activity.

**Table 4.** Sensitivity, specificity, and positive predictive value (PPV) of actigraphy and video relative to EMG. Sensitivity and specificity values for wrist actigraphy and infrared video are shown for each participant using EMG as the reference modality. Mean and standard deviation across participants are reported.

| Mount Sinai<br>RBD<br>Participant | EMG vs. Acti |  |  | EMG vs. Video |  |  |
| --- | --- | --- | --- | --- | --- | --- |
|  | Sensitivity | Specificity | PPV | Sensitivity | Specificity | PPV |
| 1 | 0.769 | 0.832 | 0.196 | 0.635 | 0.975 | 0.579 |
| 2 | 0.475 | 0.851 | 0.065 | 0.475 | 0.962 | 0.216 |
| 3 | 0.563 | 0.918 | 0.385 | 0.352 | 0.979 | 0.610 |
| 4 | 0.261 | 0.986 | 0.632 | 0.181 | 0.996 | 0.806 |
| 5 | 0.291 | 0.935 | 0.615 | 0.256 | 0.992 | 0.923 |
| 6 | 0.515 | 0.874 | 0.529 | 0.577 | 0.956 | 0.783 |
| 7 | 0.632 | 0.962 | 0.162 | 0.737 | 0.991 | 0.483 |
| 8 | 0.245 | 0.916 | 0.524 | 0.330 | 0.986 | 0.902 |
| 9 | 0.656 | 0.823 | 0.244 | 0.623 | 0.976 | 0.691 |
| Mean | 0.490 | 0.900 | 0.372 | 0.463 | 0.979 | 0.666 |
| Std Dev | 0.188 | 0.058 | 0.212 | 0.192 | 0.014 | 0.224 |

To further characterize agreement between actigraphy and video independent of EMG, we examined directional conditional probabilities between the two modalities (**Table 5**). The probability that actigraphy detected motor activity when video detected activity was high across participants (0.73 ± 0.13), indicating that most video-detected events were also captured by actigraphy. In contrast, the probability that video detected activity when actigraphy detected activity was substantially lower (0.35 ± 0.12), suggesting that a large proportion of actigraphy-detected movements were not visually apparent on video.

**Table 5.**
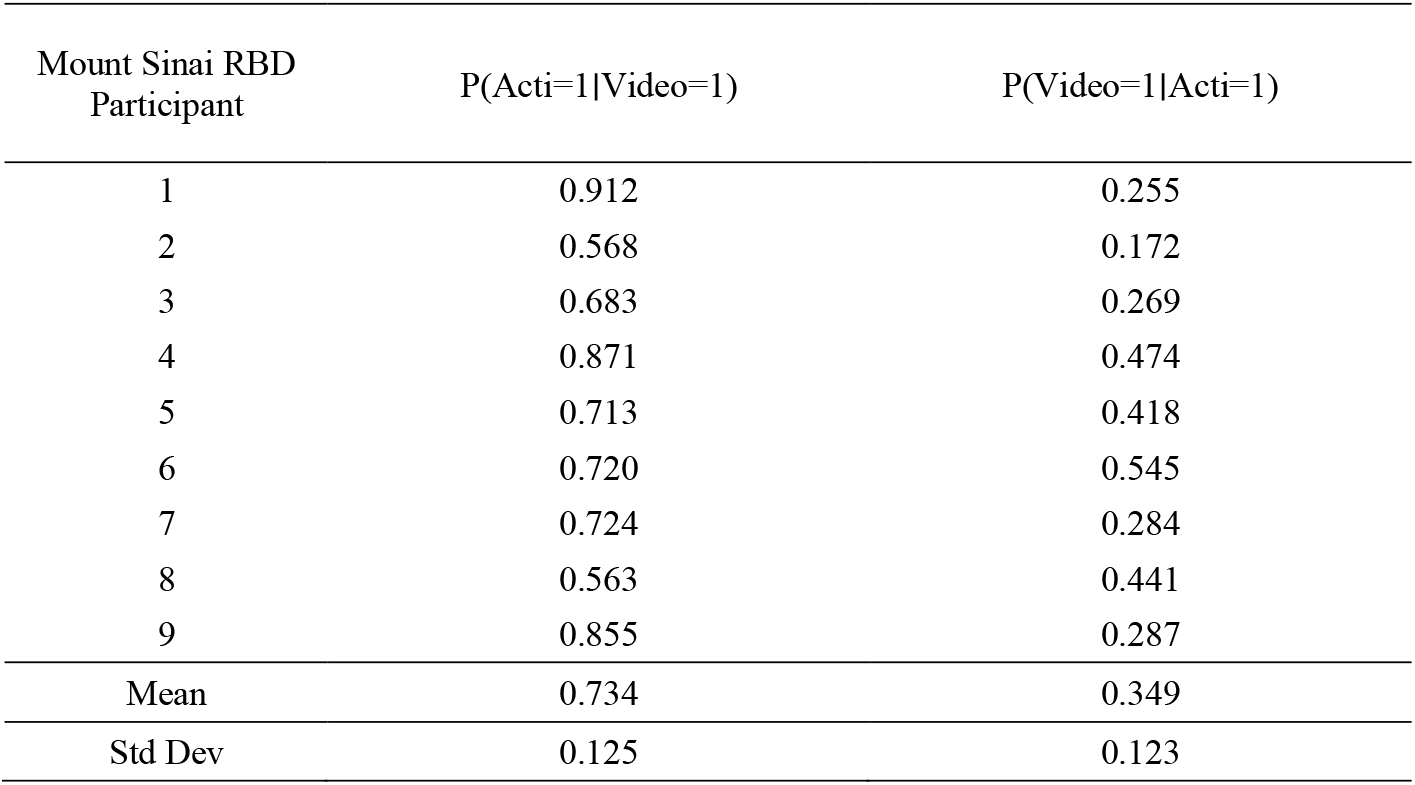
Conditional probabilities of motor activity detection across video and actigraphy. Directional conditional probabilities quantify the likelihood that motor activity detected by one modality was also detected by the other during REM sleep. Values are shown for each participant, with mean and standard deviation across participants reported.

### 3.4 REM motor activity ratio and rate measured across modalities

To compare the overall burden of REM-related motor activity detected by each modality, we measured an “activity ratio” and an “activity rate” (**Table 6**). **Activity ratio** is defined as the percentage of REM sleep mini-epochs that contain any detected arm activity. **Activity rate** is defined as the number of distinct motor events per hour of REM sleep. A motor event is counted once regardless of its duration: events may be brief (lasting <3 seconds and confined to a single mini-epoch) or prolonged (extending across multiple consecutive 3-second mini-epochs) but are considered a single event from onset to offset.

**Table 6.** Bilateral REM-related motor activity burden across EMG, actigraphy, and video during REM sleep. The proportion of REM sleep epochs containing motor activity on either left or right side (ratio, %) and the corresponding activity rate (events per hour) for each modality was computed for each participant. Mean and standard deviation across participants are reported.

| Mount Sinai<br>RBD Participant | Activity Ratio (%) |  |  | Activity Rate (events / hour) |  |  |
| --- | --- | --- | --- | --- | --- | --- |
|  | EMG | Acti | Video | EMG | Acti | Video |
| 1 | 5.04 | 19.79 | 5.53 | 32.59 | 72.16 | 32.59 |
| 2 | 2.14 | 15.56 | 4.71 | 22.46 | 106.52 | 46.20 |
| 3 | 8.34 | 12.22 | 4.82 | 57.81 | 88.84 | 29.61 |
| 4 | 8.26 | 3.41 | 1.86 | 60.32 | 26.57 | 13.64 |
| 5 | 26.37 | 12.47 | 7.31 | 133.67 | 74.20 | 42.31 |
| 6 | 21.56 | 21.00 | 15.89 | 114.67 | 101.33 | 78.67 |
| 7 | 1.14 | 4.43 | 1.74 | 8.62 | 17.25 | 9.34 |
| 8 | 27.43 | 12.82 | 10.04 | 128.01 | 52.71 | 65.08 |
| 9 | 8.03 | 21.58 | 7.24 | 42.63 | 90.00 | 31.58 |
| Mean | 12.03 | 13.70 | 6.57 | 66.76 | 69.95 | 38.78 |
| Std Dev | 10.26 | 6.63 | 4.37 | 47.09 | 31.74 | 22.42 |

Across participants, actigraphy detected the highest overall motor activity burden by both metrics, with a mean activity ratio of 13.7 ± 6.6% and a rate of 70.0 ± 31.7 per hour. EMG detected activity during a similar proportion of REM epochs (12.0 ± 10.3%) with a comparable rate (66.8 ± 47.1 events per hour), whereas video detected activity during a smaller proportion of epochs (6.6 ± 4.4%) and at substantially lower rates (38.8 ± 22.4 events per hour). Activity rates showed high inter-individual variability.

### 3.5 Actigraphy activity counts across overlapping detection conditions

Actigraphy activity counts were highest for epochs in which motor activity was detected by all three modalities (median 0.508 [IQR 0.402]) (**Figure 5**). Intermediate activity levels were observed for epochs detected by actigraphy–video pairs (median 0.197 [IQR 0.164)] and EMG–actigraphy pairs (median 0.0740 [IQR 0.141]). The lowest activity counts were observed for actigraphy-only epochs (median = 0.0958 [IQR 0.0511]).

**Figure 5.**
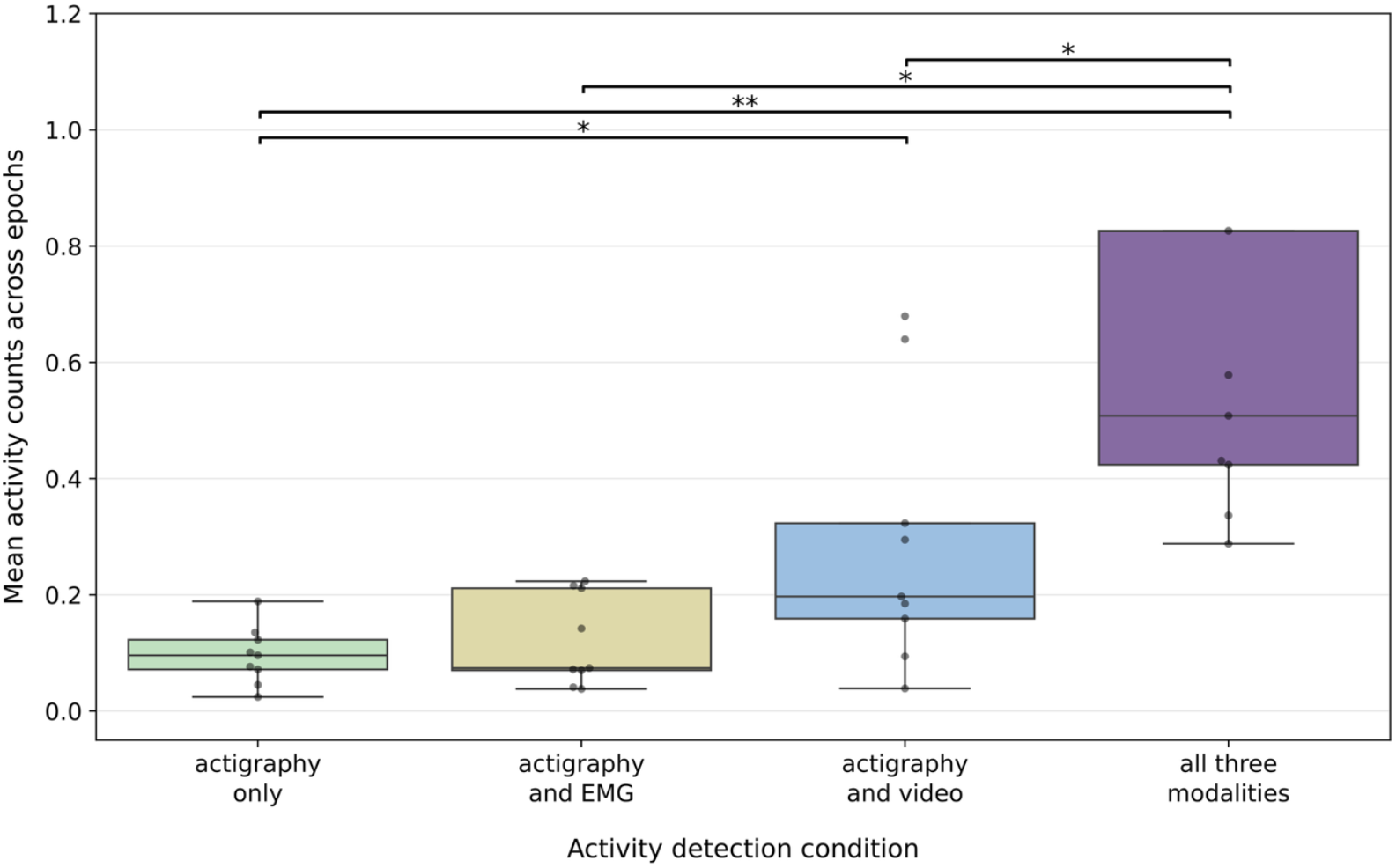
Distribution of mean actigraphy activity counts during REM sleep epochs stratified by multimodal detection condition: epochs detected by all three modalities (EMG, actigraphy, and video), by actigraphy and EMG, by actigraphy and video, or by actigraphy alone. Each dot represents a single Mount Sinai RBD participant. Actigraphy activity counts were highest for epochs detected by all three modalities and lowest for actigraphy-only epochs. Pairwise comparisons were performed using Welch’s t-test; asterisks denote statistically significant differences (*p* < 0.05). *Note*. Two values (1.53 and 1.96) in the *all three modalities* condition are not shown to improve scale and readability.

Pairwise Welch’s t-tests demonstrated that actigraphy activity counts for all-three detections were significantly greater than those for EMG–actigraphy and actigraphy-only epochs, while differences between EMG–actigraphy and actigraphy–video conditions were not statistically significant. Together, these findings indicate that higher-amplitude actigraphy signals are more likely to correspond to motor events detected by multiple modalities, whereas lower-amplitude actigraphy signals are more frequently detected in isolation.

## Discussion

In this study, we directly compared EMG, wrist actigraphy, and infrared video within synchronized REM sleep epochs to characterize how each modality captures motor activity in patients with RBD. Actigraphy-derived movement load was significantly higher during REM sleep compared to NREM stages, a pattern that was not observed in control participants. This REM-specific elevation aligns with the loss of REM atonia characteristic of RBD and demonstrates that actigraphy can capture disease-relevant motor activity during REM sleep.

Overall, agreement between video and the other two modalities—EMG (κ = 0.45 ± 0.15) and actigraphy (κ = 0.41 ± 0.12)—was highest, which is consistent with the fact that movements visible on video tend to be larger motor events that are more likely to involve both muscle activation and limb displacement. Conversely, disagreement between video and the other modalities may reflect limitations of visual detection, as some movements were obscured by bedding or body position, or occurred with minimal visible displacement despite underlying muscle activation or acceleration.

Our findings suggest that actigraphy demonstrates moderate sensitivity for detecting EMG-defined motor activity, capturing approximately half (49 ± 18.8 %) of EMG-identified motor events. Actigraphy additionally identified motor activity during epochs without concurrent EMG activation (63% of actigraphy-detected activity), consistent with its sensitivity to movements that may not be captured by the recorded EMG sensors. Importantly, actigraphy exhibited high specificity relative to EMG (90 ± 5.8 %), indicating that epochs without actigraphy-detected movement were most often associated with the absence of EMG-defined motor activity, showing its reliability in identifying periods of motor inactivity. Actigraphy’s heightened sensitivity was further reflected by it yielding the highest overall motor activity ratios (13.7 ± 6.6 %) and rates (70.0 ± 31.7 events per hour) among the three modalities.

Further analysis showed that actigraphy activity counts were significantly highest (median 0.508 [IQR 0.402]) for epochs in which motor activity was detected concurrently by all three modalities and lowest (median = 0.0958 [IQR 0.0511]) for actigraphy-only epochs, indicating that activity with higher activity counts is more likely to reflect larger, overt, multimodally detectable movements. In contrast, lower activity counts were frequently detected in isolation, suggesting that actigraphy signal magnitude may serve as a useful proxy for movement salience. These findings have practical implications for actigraphy-based RBD detection algorithms, including the potential utility of incorporating amplitude-based thresholds to improve specificity for clinically meaningful motor events.

Several limitations should be considered when interpreting these results. First, the sample (Mount Sinai cohort) size was small and included only male participants, which may limit generalizability. Second, the control group wore a different type of device, which may introduce calibration or sensitivity differences that complicate comparisons with the RBD participants. Third, motor activity was scored using binary criteria within 3-second mini-epochs, which does not capture individual movements’ duration or complexity. Video scoring was performed manually by a single rater, which may introduce human error and limits assessment of inter-rater reliability. In addition, all visible arm movements were treated equivalently without differentiation by amplitude or kinematic features, limiting more detailed characterization of video-detected activity. Finally, analyses of EMG recordings were restricted to the flexor digitorum superficialis to enable direct one-to-one comparisons with wrist actigraphy, whereas clinical vPSG assessments of RBD typically incorporate both chin and limb EMG channels for diagnosis.

Despite these limitations, our work provides a quantitative comparison of EMG, actigraphy, and video within the same REM sleep periods, extending prior RBD diagnostic and actigraphy studies to the level of motor-event concordance across modalities. This framework provides clinicians with a quantitative understanding of the complementary strengths of EMG, video, and actigraphy, and informs the potential role of actigraphy in scalable RBD screening or longitudinal monitoring outside of the laboratory. Future work should extend these analyses to larger and more diverse cohorts. Additional studies should compare EMG with actigraphy across various body placements (wrist, trunk, ankles) and 3D time-of-flight video, which is more sensitive than conventional infrared video systems^5^.

## Data Availability

Control data used in this study is openly available in Zenodo at 10.5281/zenodo.1160410. RBD participant data are available upon reasonable request to the authors.

https://zenodo.org/records/1160410

## Author Contributions

Conceptualization, KHR, GRM, and EHD; Methodology, KHR, GRM, SM, ABK, and EHD.; Software, KHR, GRM, and ABK.; Validation, KHR, GRM, ABK, and EHD; Formal Analysis, KHR, GRM, and ABK; Investigation, KHR, GRM, SM, and EHD; Resources, ABK and EHD; Data Curation, KHR, SM, and EHD; Writing – Original Draft Preparation, KHR and EHD; Writing – Review & Editing, KHR, GRM, SM, ABK, and EHD; Visualization, KHR and GRM; Supervision, EHD; Project Administration, EHD; Funding Acquisition, EHD.

## Data availability

Control data used in this study is openly available in Zenodo at 10.5281/zenodo.1160410. RBD participant data are available upon reasonable request from the authors.

## Conflicts of Interest

The authors declare no conflict of interest.

## Informed Consent

Informed consent for participation was obtained from all subjects involved in the study.

## Institutional Review Board Statement

The study was conducted in accordance with the Declaration of Helsinki, and approved by the Institutional Review Boards of the Icahn School of Medicine at Mount Sinai (protocol code 22-00977 and date of approval Nov 1 2022).

## Funding Sources

Department of Neurology, Icahn School of Medicine at Mount Sinai

## Acknowledgements

We thank all study participants for enabling this research and the Department of Neurology at the Icahn School of Medicine for funding.

## Notes

### Competing Interest Statement

The authors have declared no competing interest.

### Funding Statement

This study was funded by the Department of Neurology, Icahn School of Medicine at Mount Sinai.

### Author Declarations

The Institutional Review Boards of the Icahn School of Medicine at Mount Sinai gave ethical approval for this work.

