## Supplementary Materials for "Comparison of EMG, Video, and Actigraphy Signals for Detecting Motor Activity in REM Sleep Behavior Disorder"

| Stanford RBD Participant | Total sleep time (minutes) | Total REM duration (minutes) | REM sleep periods (number) |
| --- | --- | --- | --- |
| 1 | 415 | 121 (29.2%) | 3 |
| 2 | 361.5 | 66 (18.3%) | 4 |
| 3 | 400.5 | 52.5 (13.1%) | 4 |
| 4 | 502.5 | 206.5 (41.1%) | 3 |
| 5 | 408 | 140 (34.3%) | 4 |
| 6 | 418.5 | 76.5 (18.3%) | 2 |
| 7 | 210 | 47 (22.4%) | 3 |
| 8 | 484 | 53.5 (11.1%) | 3 |

**Table S1.** Sleep architecture characteristics for each RBD participant in the Stanford cohort. Total sleep time, total REM sleep duration (with percentage of total sleep time), and number of REM sleep periods during the recorded overnight video-polysomnography (vPSG) are shown for each participant.

| Newcastle Control Participant | Total sleep time (minutes) | Total REM duration (minutes) | REM sleep periods (number) |
| --- | --- | --- | --- |
| 1 | 427.5 | 61 (14.3%) | 2 |
| 2 | 507.5 | 21.5 (4.24%) | 2 |
| 3 | 316.5 | 45 (14.2%) | 3 |
| 4 | 563.5 | 143.5 (25.5%) | 4 |
| 5 | 464.5 | 75.5 (16.3%) | 3 |
| 6 | 248.5 | 19.5 (7.85%) | 1 |
| 7 | 483 | 113.5 (23.5%) | 3 |
| 8 | 490.5 | 158.5 (32.3%) | 4 |
| 9 | 445 | 88 (19.8%) | 4 |

**Table S2.** Sleep architecture characteristics for each control participant in the Newcastle open dataset. Total sleep time, total REM sleep duration (with percentage of total sleep time), and number of REM sleep periods during the recorded overnight video-polysomnography (vPSG) are shown for each participant.

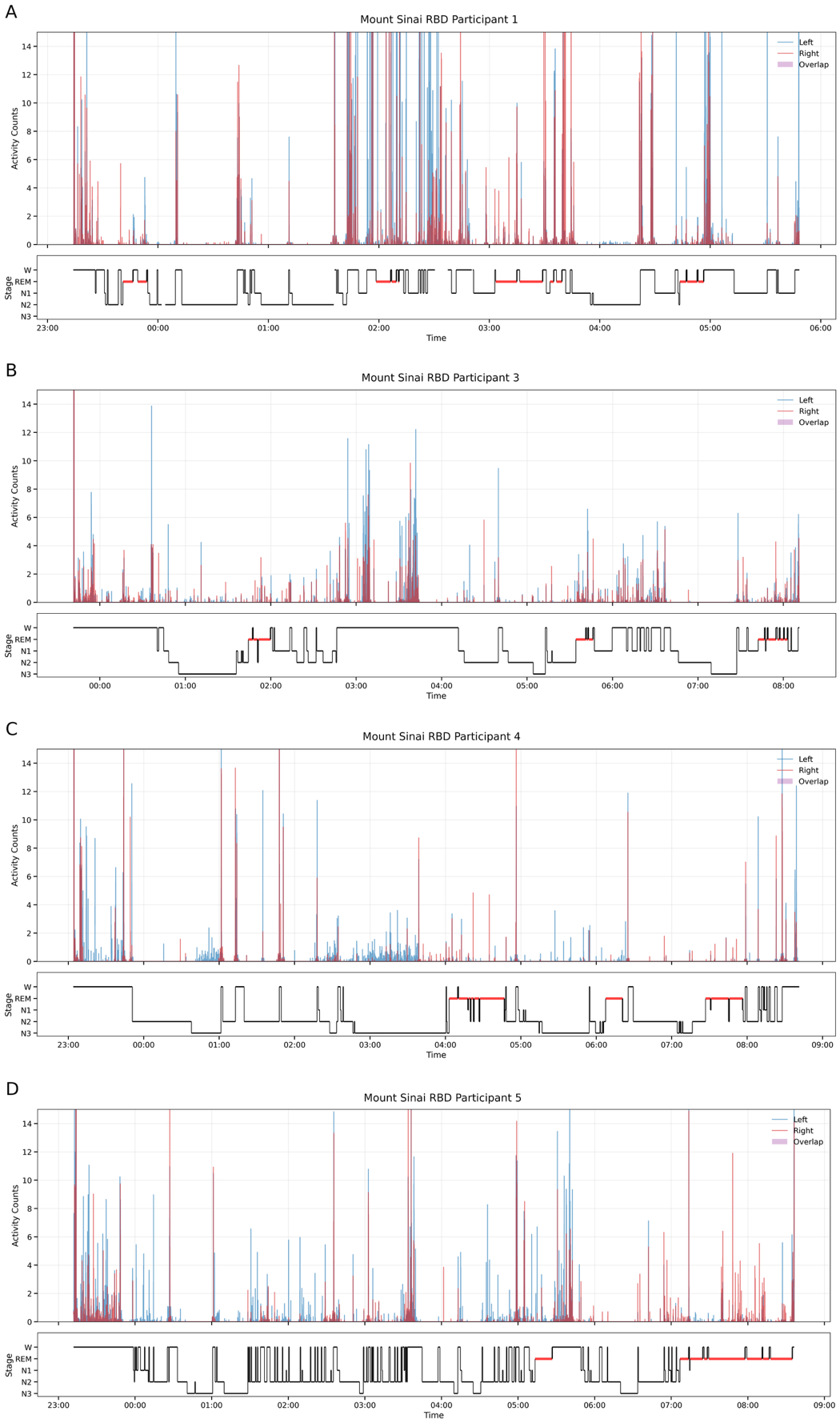

**Figure S1.** Bilateral wrist actigraphy aligned to sleep stages in Mount Sinai RBD participants (1, 3, 4, 5). Panels A-D correspond to participants 1, 3, 4, and 5, respectively. The upper panel shows left- (blue) and right-wrist (red) activity counts across the overnight recording; purple shading marks epochs with concurrent activity in both wrists (overlap). The lower panel shows the corresponding hypnogram (W, N1, N2, N3, REM), with REM periods highlighted in red. *Note.* The y-axis is truncated at 15 to improve readability.

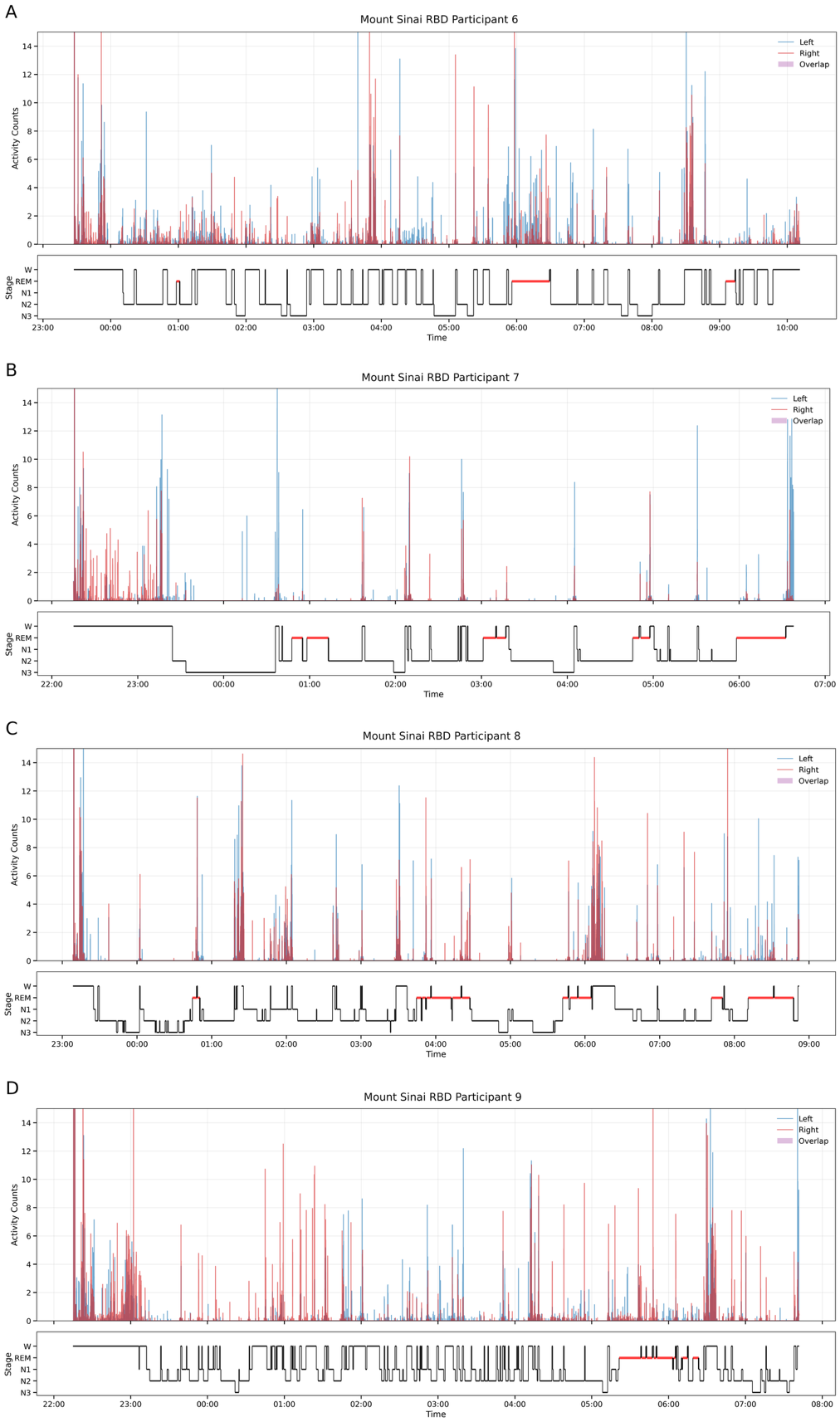

**Figure S2.** Bilateral wrist actigraphy aligned to sleep stages in Mount Sinai RBD participants (6-9). Panels A-D correspond to participants 6, 7, 8, and 9, respectively. The upper panel shows left- (blue) and right-wrist (red) activity counts across the overnight recording; purple shading marks epochs with concurrent activity in both wrists (overlap). The lower panel shows the corresponding hypnogram (W, N1, N2, N3, REM), with REM periods highlighted in red. *Note.* The y-axis is truncated at 15 to improve readability.

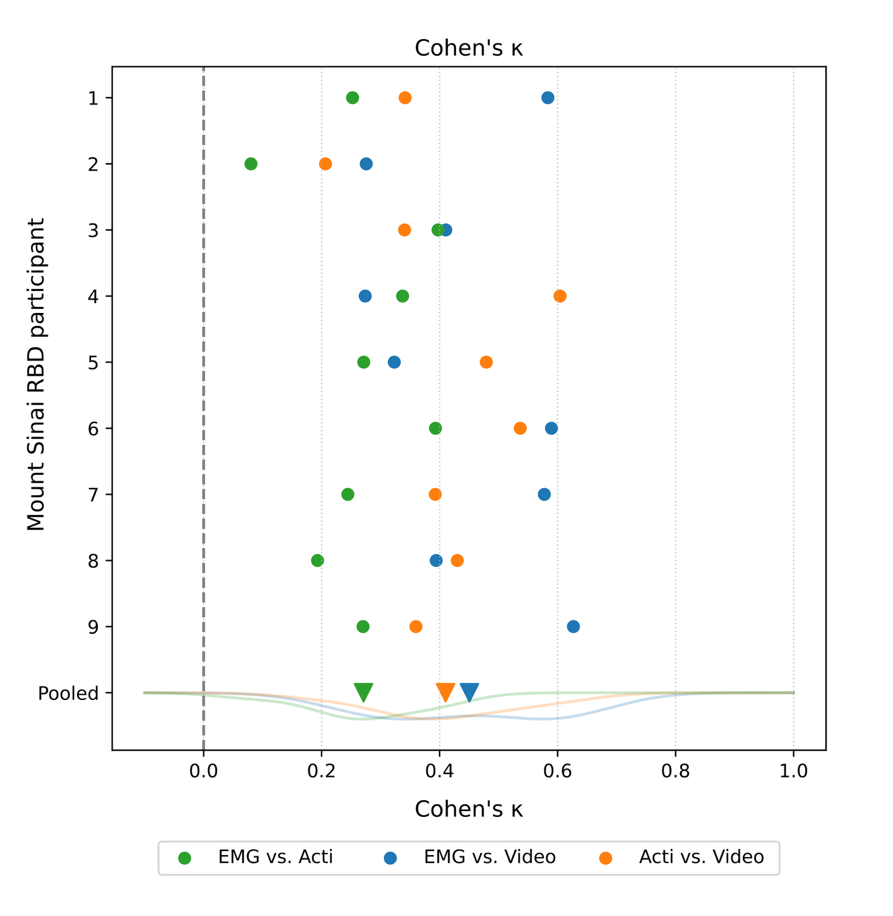

**Figure S3.** Agreement between EMG, actigraphy, and video detections during REM sleep. Cohen’s κ is shown for pairwise comparisons between EMG and actigraphy (green), EMG and video (blue), and actigraphy and video (orange) for each participant. The downward triangles denote the pooled mean κ across participants for each comparison.

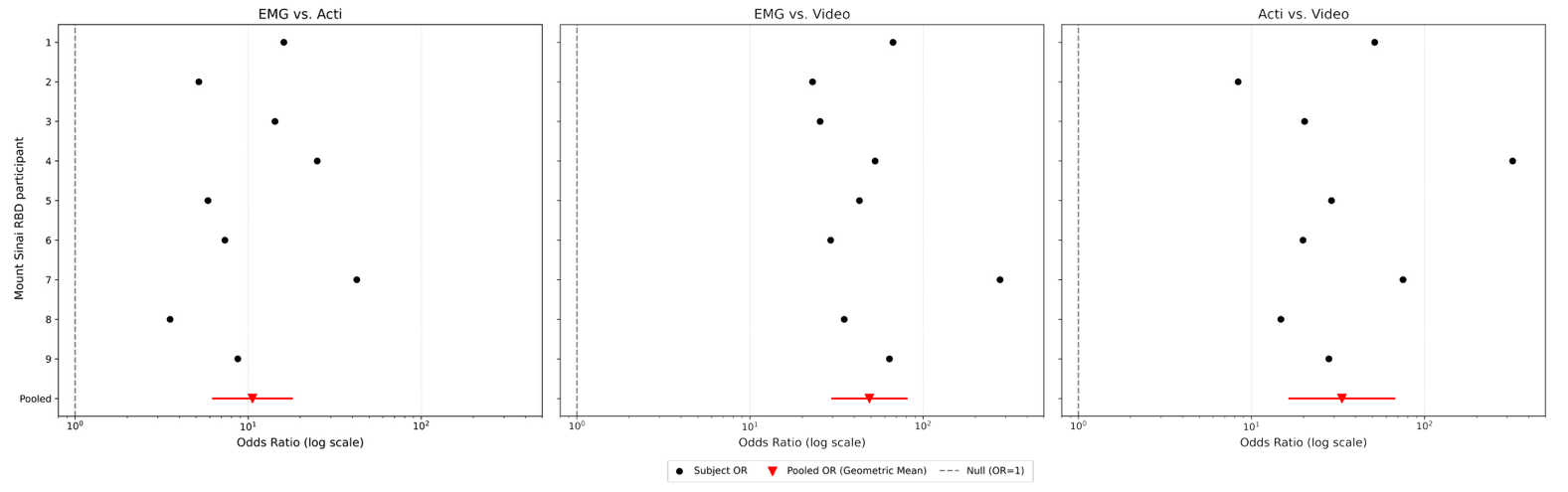

**Figure S4.** Pairwise association between EMG, actigraphy, and video detections during REM sleep. Odds ratios (ORs) are shown on a log scale for each Mount Sinai RBD participant (black dots) for EMG vs. actigraphy (left), EMG vs. video (middle), and actigraphy vs. video (right). The pooled OR across participants is shown in red (downward triangle) with its confidence interval (red horizontal line). The dashed vertical line indicates the null value (OR = 1).
